# First results from the UK COVID-19 Serology in Oncology Staff Study (CSOS)

**DOI:** 10.1101/2020.06.22.20136838

**Authors:** DM Favara, A Cooke, R Doffinger, S Houghton, I Budriunaite, S Bossingham, K McAdam, P Corrie, N Ainsworth

## Abstract

**Background:** The global SARS-CoV-2 (COVID-19) pandemic has caused substantial worldwide mortality. At present, there is no data regarding oncologist-specific SARS-CoV-2 infection/immunity rates in the United Kingdom (UK) which might impact planning for the management of potentially immunosuppressed cancer patients. Here, we present the first results from the COVID-19 Serology in Oncology Staff (CSOS) study with the aim of informing non-surgical oncology management guidelines.

**Methods:** Patient-facing staff working in an oncology department during the COVID-19 pandemic at a large district general hospital in the East of England were invited to participate. Samples were collected during the first week of June 2020: blood for SARS-COV-2 antibody testing using a rapid lateral flow point of care (POC) assay and a laboratory Luminex based assay, as well as a nasopharyngeal swab for SARS-CoV-2 PCR testing. Participant characteristics were also collected.

**Results:** Seventy participants were recruited: nurses (45/70; 64.3%), doctors (15/70; 21.2%), and other patient-facing staff (10/70; 14.3%). The majority were female (61/70; 87.1%) with a mean age of 42 years (median 41; range 23-64 years). A minority were smokers (9/70; 10%) or had chronic underlying health conditions (16/70; 22.9%), the commonest being asthma. All participants were nasopharyngeal-swab PCR negative, although 4/70 (5.7%) had previously tested positive by NHS testing undertaken during the preceding months. 15/70 (21.4%) had positive SARS-CoV-2 antibodies using the Luminex test. Nurses had the highest incidence of positive antibodies (13/45; 28.9%), with a lower incidence in doctors (2/15; 13.3%) although this difference was not statistically significant (Fischer’s exact test *p*=0.3). No receptionists had positive antibody tests. All four participants with a previously reported positive PCR test were antibody-positive. 9/15 (60%) of antibody-positive participants reported previous symptoms suggestive of SARS-CoV-2 infection: a 3.6-fold higher odds than antibody-negative participants, of whom 16/55 reported symptoms (*p*=0.03). The mean duration of symptoms was 11 days (median 11; range 1-35 days) and the mean time from resolution of reported previous symptoms to antibody testing was 48.4 days (median 46; range 1-123 days).

**Conclusion:** This study establishes the SARS-CoV-2 exposure and carriage rate amongst patient-facing staff working in the oncology department of a large UK general hospital during the pandemic. These results may help inform UK national oncology patient management prior to the development of a viable vaccine or treatment.

## Introduction

The global SARS-CoV-2 (COVID-19) pandemic has caused substantial morbidity and economic turmoil across the world. Despite ongoing research endeavours, SARS-CoV-2-related deaths continue to rise globally without any clear sight of an effective treatment, vaccine or virus eradication. Various populations in multiple countries have been tested for SARS-CoV-2 infection using PCR to detect viral antigen, with a wide range of reported asymptomatic positive carrier rates (4-80%).^1–5^ Antibody tests are the gold standard for providing evidence of exposure to pathogens following an adaptive immune response, including SARS-CoV-2 infection.^6,7^ Although reports of IgM and IgG antibody production rates against SARS-CoV-2 in COVID-19 pandemic survivors has varied (as reported in the media), the first large US series (n=285) of hospitalised SARS-CoV-2 PCR positive patients reported that 100% of them developed SARS-CoV-2 IgG within 19 days of symptom onset.^8^ Another US study showed that 511/624 (82%) of SARS-CoV-2 PCR positive out-patients had developed antibodies 10-14 days after symptom onset.^9^ In a French study, 10/14 (71%) SARS-CoV-2 PCR positive Health Care Workers (HCWs) developed detectable SARS-CoV-2 antibodies 15 days after PCR positivity and symptom onset.^10^

The percentage of asymptomatic HCWs who are SARS-CoV-2 IgG positive (without working solely in a SARS-CoV-2 ward) has not been extensively investigated and may be a useful indicator of population-wide infection rates during nationwide lockdown. Current data suggests that this may be very low, as evidenced by a French study in which 3/230 (1.3%) asymptomatic PCR-naïve HCWs were SARS-CoV-2 IgG-positive, with all 3 being PCR negative at the time of testing, suggesting past exposure with successful eradication of the virus.^10^ A Spanish study from a tertiary hospital reported a similarly low SARS-CoV-2 antibody-positive rate in asymptomatic staff (11/578; 1.9%).^11^ Data from screening asymptomatic HCWs for active SARS-CoV-2 viral infection is limited. One United Kingdom (UK) study showed a peak asymptomatic HCW infection at 7.1% in late March 2020, decreasing to 1.1% five weeks later^12^, whilst another UK study showed that only 30 of 1032 (3%) asymptomatic HCWs tested at a tertiary hospital in April 2020 were SARS-CoV-2 PCR positive.^13^

People with cancer may be more likely to contract SARS-CoV-2 infection.^14,15^ This risk is amplified by multiple hospital attendances required for diagnosis, treatment and follow-up. Although recent results suggest that anti-cancer treatment does not increase mortality in infected cancer patients^16^, it may increase the risk of serious complications following infection^14,15^, so guidance is needed to safeguard both patients and oncology staff throughout their treatment pathways. There is currently no data regarding oncologist-specific SARS-CoV-2 infection and immunity rates within the UK, while risk of viral transmission between HCWs working with cancer patients is not known. The ‘COVID-19 Serology in Oncology Staff’ (CSOS) study is a multi-centre UK study aiming to provide an indication of staff infection rates in order to influence UK national guidance in managing cancer patients treated within secondary care oncology departments, which is currently lacking. The CSOS study investigates SARS-CoV-2 serology (IgG) and asymptomatic viral carriage within patient-facing oncology staff 10 weeks after the UK’s COVID-19 pandemic national lockdown, with sample collection at multiple time points. The primary objective is to measure the percentage of SARS-CoV-2 IgG positive asymptomatic oncology staff in the workplace during the pandemic. Secondary outcomes include the proportion of previously symptomatic and asymptomatic SARS-CoV-2 IgG seropositivity, the rate of persistent asymptomatic PCR positivity over time, and the proportion of those who do not become antibody-positive following a positive PCR result. Here, we present the first study findings from testing staff in the oncology department at a single, large district general hospital in the East of England.

## Methods

We recruited staff involved in treating both oncology and haemato-oncology patients in the oncology department at The Queen Elizabeth Hospital in Kings Lynn (QEHKL), a 515-bed district general hospital in the East of England serving a population of approximately 331,000 people. Staff were invited to participate in the study unless they had not been at work at all for the duration of the pandemic, or were not patient-facing. Participants had to have been working within the oncology department and not primarily within a dedicated SARS-CoV-2 in-patient ward. Individuals who returned to work after self-isolating due to SARS-CoV-2 symptoms, or exposure to a potentially affected household member (as per UK government rules) were eligible to participate. Following consent, samples were collected during the first week of June 2020, specifically, blood for SARS-COV-2 antibody testing and a nasopharyngeal swab for SARS-CoV-2 PCR testing. The following anonymised participant characteristics were collected: age, sex, role, smoking status, details of suspected SARS-CoV-2 illness/exposure and leave taken, dates of start to resolution of presumed or confirmed SARS-CoV-2 illness, date of SARS-CoV-2 tests and results, and history of any underlying health condition. Ethical approval for the study was granted by the United Kingdom’s Health Research Authority (HRA) (IRAS: 284231; 26/5/2020). Hospital approval was granted by the Queen Elizabeth Hospital King’s Lynn NHS Foundation Trust R&D Department (Ref: 9/20; 28/5/2020). Data was analysed using Prism 8 (Graphpad Software). Fischer’s exact test and an unpaired Student’s *t*-test were used.

### SARS-CoV-2 serology

Two different antibody assays were used to detect SARS-CoV-2 antibodies: a rapid point of care (POC) assay and a laboratory-based assay. The rapid POC test used was the Abbexa (Cambridge, United Kingdom) COVID-19 IgG/IgM Rapid Test Kit (abx294171) detecting antibodies against the SARS-CoV-2 nucleocapsid (N) and spike (S) proteins. Blood was collected from a finger prick as per manufacturer specifications. The manufacturer claims that it has no cross-reactivity with antibodies against other coronavirus types (HKU1, OC43, NL63, 229E) and that it has a sensitivity of 98.5% and a specificity of 97.94%. Results were read 10-15 minutes after assaying.

The laboratory-based assay used was a SARS-CoV-2 IgG multiplex particle-based flow cytometry (Luminex) assay developed at Cambridge University Hospitals NHS Foundation Trust (Cambridge, United Kingdom) detecting antibodies against the SARS-CoV-2 N and S proteins. Approximately 4mL of blood was collected in a serum tube for this assay. Its sensitivity and specificity were determined at 84% and 100% respectively, using a cohort of pre-pandemic healthy controls versus a cohort of unselected SARS-CoV2 PCR-positive patients. At time of writing, testing for cross-reactivity against other coronaviruses had not been performed for this semi-quantitative assay.

### SARS-CoV-2 PCR

RNA extraction was performed using the Zymo (Cambridge, UK) Quick-RNA 96 kit (R1053). SARS-CoV-2 detection by PCR was performed using the Primerdesign (Eastleigh, UK) Coronavirus COVID-19 Genesig RT-PCR assay (Z-Path-COVID-19-CE) which included positive and internal extraction controls. All kits were used according to manufacturer guidelines. qPCR was run using Roche LightCycler 480 in a 96-well format.

## Results

Seventy staff out of total of 82 eligible oncology department staff were recruited to the study (85.4% participation rate). Samples were collected during the first week of June 2020. Participant characteristics are shown in **Table 1**. The majority of participants were nurses (45/70; 64.3%), followed by doctors (15/70; 21.2%), and other patient-facing staff (10/70; 14.3%). The majority of participants were female (61/70; 87.1%). The mean participant age was 42 years (median 41; range 23-64 years) with a similar mean (and median) for both doctors and nurses. Only 7/70 (10%) of participants were smokers: nurses (5/45; 11.1%), receptionists (2/10, 20%). Chronic underlying health conditions were reported in 16/70 (22.9%) of all participants, with asthma (5/70; 7.1%) and hypertension (4/70; 5.7%) being the most common for all groups and receptionists having the highest prevalence (5/10; 50%). From the start of the COVID-19 pandemic to the sample collection date (March – June 2020), 9 cases of PCR-confirmed SARS-CoV-2 infection had been recorded in cancer patients treated on the QEHKL oncology in-patient ward out of a total of 150 in-patient admissions during this period (9/150; 6%).

**Table 1:** participant characteristics

|  | Total | Doctors | Nurses | Receptionists |
| --- | --- | --- | --- | --- |
| **Number** | 70 | 15 | 45 | 10 |
| **Mean age (median; range)** | 42 (41; 23-64) | 41 (42; 25-60) | 40 (40; 23-61) | 48 (49; 25-64) |
| **Female** | 61/70 (87.1%) | 9/15 (60%) | 42/45 (93.3%) | 10/10 (100%) |
| **Male** | 9/70 (12.9%) | 6/15 (40%) | 3/45 (6.7%) | 0/10 (%) |
| **Smoker** | 7/70 (10%) | 0/15 (0%) | 5/45 (11.1%) | 2/10 (20%) |
| **Underlying health condition** | 16/70 (22.9%) | 2/15 (13.3%) | 9/45 (20%) | 5/10 (50%) |
| *asthma* | 5/70 (7.1%) | 1/15 (6.7%) | 3/45 (6.7%) | 1/10 (10%) |
| *hypertension* | 4/70 (5.7%) | 0/15 (0%) | 2/45 (4.4%) | 2/10 (20%) |
| *diabetes* | 1/70 (1.4%) | 0/15 (0%) | 1/45 (2.2%) | 0/10 (0%) |
| *hyperthyroidism* | 1/70 (1.4%) | 1/15 (6.7%) | 0/45 (0%) | 0/10 (0%) |
| *anaemia* | 1/70 (1.4%) | 0/15 (0%) | 1/45 (2.2%) | 0/10 (0%) |
| *thrombocytopaenia* | 1/70 (1.4%) | 0/15 (0%) | 1/45 (2.2%) | 0/10 (0%) |
| *COPD* | 1/70 (1.4%) | 0/15 (0%) | 1/45 (2.2%) | 0/10 (0%) |
| *cardiac disease* | 1/70 (1.4%) | 0/15 (0%) | 0/45 (0%) | 1/10 (10%) |
| *previous cancer* | 1/70 (1.4%) | 0/15 (0%) | 0/45 (0%) | 1/10 (10%) |

Results (**Table 2**) showed that 25/70 (35.7%) of participants reported prior symptoms of SARS-CoV-2 infection, with nurses having the highest incidence (17/45; 37.7%) and receptionists having the lowest (3/10; 30%). Mean duration of reported symptoms was 11 days (median 11; range 1-35 days). Duration was similar for all staff groups. Of those with reported previous symptoms, 11/25 (44%) underwent PCR nasopharyngeal testing for SARS-CoV-2 infection at the time; 4/11 (36.4%) tested positive. Only 5/17 (29.4%) previously symptomatic nurses received a prior PCR test (2/5; 40% were positive), in contrast to (4/5; 80%) previously symptomatic doctors were tested (2/4; 50% were positive). The mean time from resolution of reported previous symptoms to the CSOS study sample collection date was 48.4 days (95% CI 39.3-57.46). 45/70 (64.3%) participants reported no prior symptoms during the pandemic, which was similar across all groups.

**Table 2:**
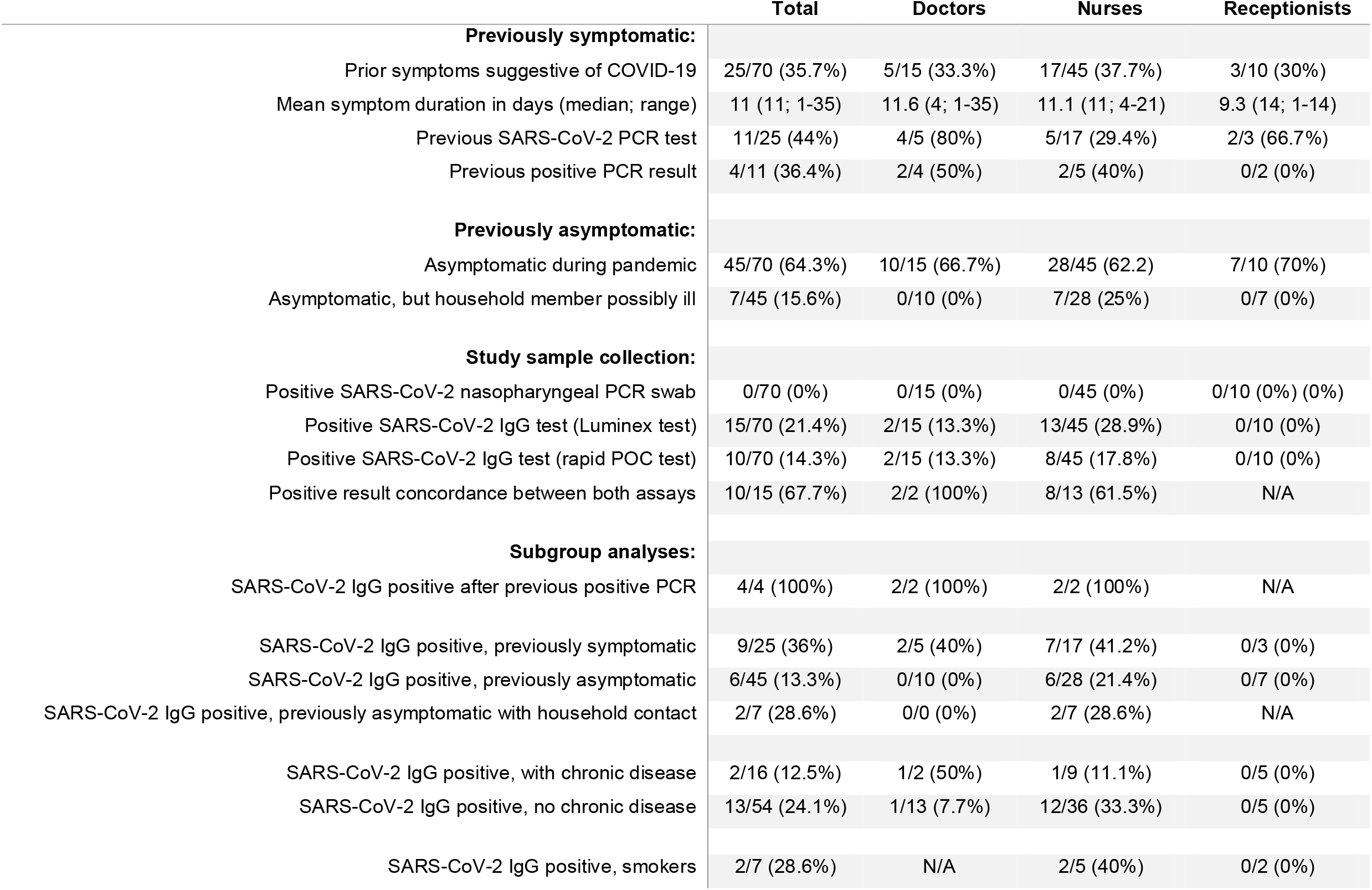
Results

|  | Total | Doctors | Nurses | Receptionists |
| --- | --- | --- | --- | --- |
| **Previously symptomatic:** |  |  |  |  |
| Prior symptoms suggestive of COVID-19 | 25/70 (35.7%) | 5/15 (33.3%) | 17/45 (37.7%) | 3/10 (30%) |
| Mean symptom duration in days (median; range) | 11 (11; 1-35) | 11.6 (4; 1-35) | 11.1 (11; 4-21) | 9.3 (14; 1-14) |
| Previous SARS-CoV-2 PCR test | 11/25 (44%) | 4/5 (80%) | 5/17 (29.4%) | 2/3 (66.7%) |
| Previous positive PCR result | 4/11 (36.4%) | 2/4 (50%) | 2/5 (40%) | 0/2 (0%) |
| **Previously asymptomatic:** |  |  |  |  |
| Asymptomatic during pandemic | 45/70 (64.3%) | 10/15 (66.7%) | 28/45 (62.2%) | 7/10 (70%) |
| Asymptomatic, but household member possibly ill | 7/45 (15.6%) | 0/10 (0%) | 7/28 (25%) | 0/7 (0%) |
| **Study sample collection:** |  |  |  |  |
| Positive SARS-CoV-2 nasopharyngeal PCR swab | 0/70 (0%) | 0/15 (0%) | 0/45 (0%) | 0/10 (0%) (0%) |
| Positive SARS-CoV-2 IgG test (Luminex test) | 15/70 (21.4%) | 2/15 (13.3%) | 13/45 (28.9%) | 0/10 (0%) |
| Positive SARS-CoV-2 IgG test (rapid POC test) | 10/70 (14.3%) | 2/15 (13.3%) | 8/45 (17.8%) | 0/10 (0%) |
| Positive result concordance between both assays | 10/15 (67.7%) | 2/2 (100%) | 8/13 (61.5%) | N/A |
| **Subgroup analyses:** |  |  |  |  |
| SARS-CoV-2 IgG positive after previous positive PCR | 4/4 (100%) | 2/2 (100%) | 2/2 (100%) | N/A |
| SARS-CoV-2 IgG positive, previously symptomatic | 9/25 (36%) | 2/5 (40%) | 7/17 (41.2%) | 0/3 (0%) |
| SARS-CoV-2 IgG positive, previously asymptomatic | 6/45 (13.3%) | 0/10 (0%) | 6/28 (21.4%) | 0/7 (0%) |
| SARS-CoV-2 IgG positive, previously asymptomatic with household contact | 2/7 (28.6%) | 0/0 (0%) | 2/7 (28.6%) | N/A |
| SARS-CoV-2 IgG positive, with chronic disease | 2/16 (12.5%) | 1/2 (50%) | 1/9 (11.1%) | 0/5 (0%) |
| SARS-CoV-2 IgG positive, no chronic disease | 13/54 (24.1%) | 1/13 (7.7%) | 12/36 (33.3%) | 0/5 (0%) |
| SARS-CoV-2 IgG positive, smokers | 2/7 (28.6%) | N/A | 2/5 (40%) | 0/2 (0%) |

All participants tested at day 0 of the study were nasopharyngeal swab PCR negative for SARS-CoV-2 (**Table 2**). A positive SARS-CoV-2 IgG was detected in 15/70 (21.4%) of participants using the Luminex test, and in 10/70 (14.3%) using the rapid POC serology test. All participants positive using the rapid POC serology test had also been positive using the Luminex test. Due to its ability to detect lower antibody concentration levels (because of the assay type), results from the Luminex assay were used as the final result. Nurses had the highest percentage of SARS-CoV-2 antibodies (13/45; 28.9%). The percentage prevalence in doctors was less than half of that in nurses (2/15; 13.3% although this difference was not significant (Fischer’s exact test *p*=0.3). No SARS-CoV-2 antibodies were detected in the receptionists. All participants with a positive nasopharyngeal PCR result prior to the study tested positive for antibodies (4/4; 100%).

60% (9/15) of antibody-positive participants reported previous symptoms consistent with SARS-CoV-2 infection during the pandemic: a 3.6 fold higher odds than antibody-negative participants (16/55; 29.1%) (Fischer’s exact test *p*=0.03). Out of the total number of previously symptomatic participants, 9/25 (36%) had detectable SARS-CoV-2 antibodies. In those who reported no prior symptoms during the pandemic, 6/45 (13.3%), all of whom where nurses, had antibodies, indicating asymptomatic prior infection. Out of 7 participants who had no prior symptoms but had been exposed to a suspected infected household member, 2/7 (28.6%) had positive antibodies. These findings are visualised in **Figure 1**. 2/16 (12.5%) of participants with chronic underlying health conditions had positive antibodies in contrast to 13/54 (24.1%) of those without. Of the smokers, only 2/5 (40%) were antibody-positive in the nurse group, with no positives in the receptionist group.

**Figure 1:**
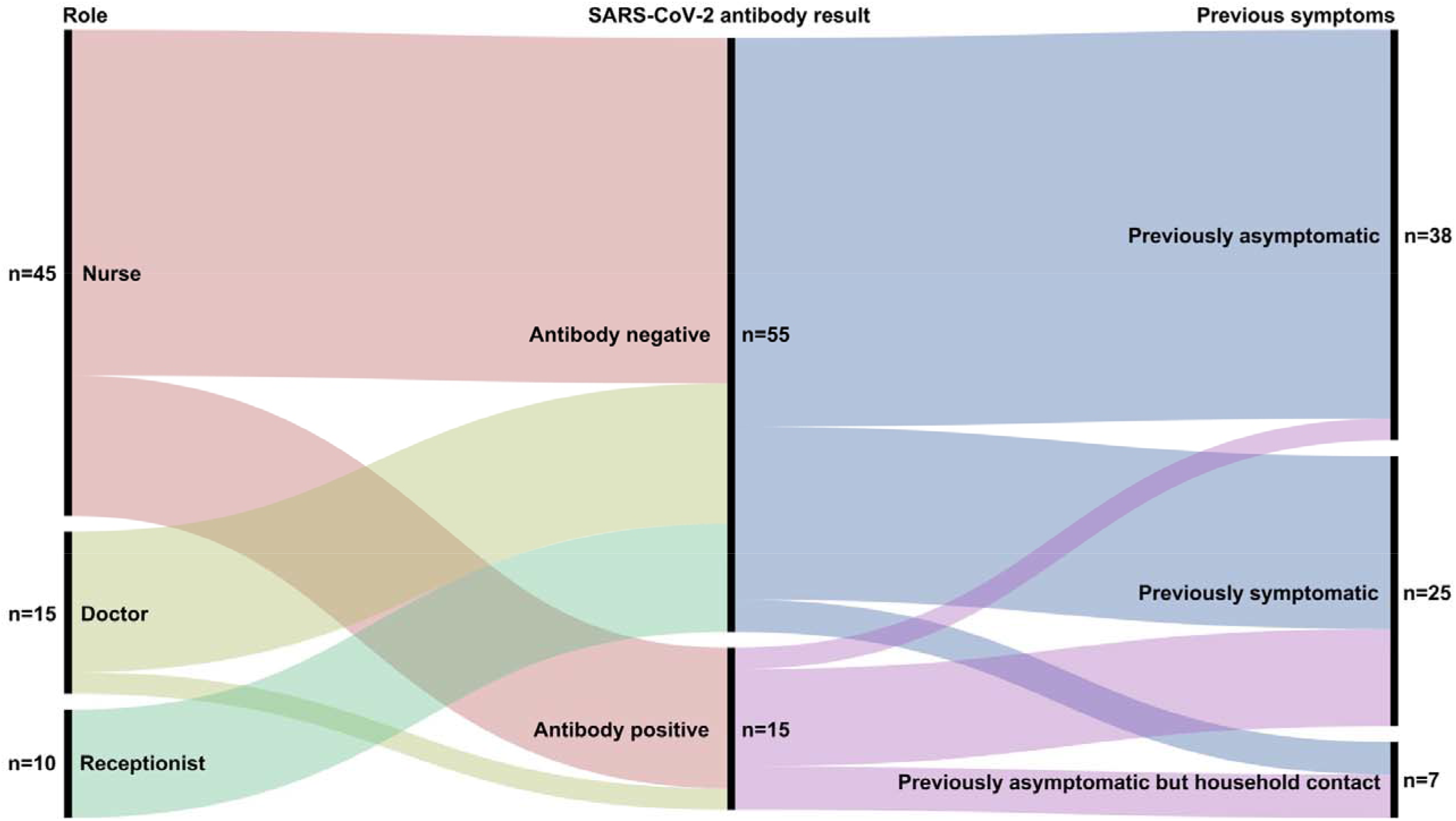
Summary of relationship between role, previous symptoms and antibody result. All participants were nasopharyngeal swab SARS-COV-2 PCR negative at time of SARS-COV-2 antibody testing.

## Discussion

To the best of our knowledge, this is the first UK study specifically investigating SARS-CoV-2 exposure in patient-facing oncology staff who were at work within a secondary care non-surgical oncology department during the COVID-19 pandemic between March and the start of June 2020. Nearly a quarter of oncology staff assessed were SARS-CoV-2 antibody-positive, suggesting a substantial past infection rate, although we found that no participants were SARS-CoV-2 PCR positive at the time of sampling. Although only 6% of the patients admitted to the oncology in-patient ward during the first 3 months of the UK lockdown were found to be PCR-positive, by the nature of the hospital admission process it is possible that some of the infections amongst staff (both previously symptomatic and asymptomatic) could have arisen from exposure to these patients, especially earlier on during the pandemic, personal protective equipment (PPE) was less readily available within the NHS.

Nurses were the staff group with the highest percentage of positive SARS-CoV-2 antibodies (double that of doctors, although this difference was not statistically significant at this sample size), which if borne out in a larger sample size, may be the result of higher frequency and duration of physical contact between nurses and patients by nature of their work. That none of the receptionist group were antibody-positive fits with this hypothesis. A higher proportion of those who reported prior symptoms suggestive of SARS-CoV-2 infection were antibody-positive. However, our finding of a 13.3% asymptomatic infection rate (evidenced by positive antibodies) is higher than reported elsewhere in HCWs^10,11^ It however remains unclear whether such antibodies are protective against future repeat SARS-CoV-2 infection.

In order to limit the possibility of erroneous results, we used two different antibody detection methods. The rapid POC antibody test was reported by the manufacturer to have high sensitivity and specificity and not to cross-react with the 4 main other coronavirus types, whilst the Luminex test was able to detect antibodies at a lower concentration level (by the nature of the method). This was evidenced by a previously SARS-CoV-2 PCR-positive participant, who was confirmed to be low level anti-SARS-CoV-2 (IgG)-positive by the Luminex test, but not by the rapid POC method. The Luminex test had not been investigated for cross reactivity against other coronaviruses (at time of writing). Hence, we cannot exclude the possibility that some of the participants who were positive using the Luminex antibody test had not actually had SARS-CoV-2 infection, but had instead had been exposed to other coronaviruses. If we had used the rapid test only, the overall positive antibody percentage would have been 8% lower. Although there is the possibility that some of our study participants were recently SARS-CoV-2 infected and thus were not yet producing SARS-CoV-2 IgG or had fully seroconverted), the mean time from the reported resolution of previous symptoms to the start of the study was 1.5 months. This is something which will be explored with additional sample collection at a later time point.

This study is ongoing and will be collecting further samples from the QEHKL cohort as well as at other NHS hospitals. We report these interim results in the expectation that they will be of importance for planning UK national guidance on SARS-CoV-2 testing of patients due to start or having started anticancer non-surgical treatments, as well as the oncology staff treating them.

## Data Availability

Data available following reasonable request.

## Acknowledgements

The authors would like to thank Sisters Mandy Coventry and Paula Santos for their help in sample collection at QEHKL, Dr Hilary Martin (Wellcome Sanger Institute and University of Cambridge) and Dr Mireia Crispin Ortuzar (CRUK Cambridge Institute, University of Cambridge) for their critical readings of the manuscript.

## Author contributions

DMF designed the study and wrote the manuscript. NA, PC, and KA contributed to critical revision of the study and manuscript and read and approved the final version. RD, SH, IB and SB processed the Luminex SARS-CoV-2 assays, and AC processed the SARS-CoV-2 PCR assays.

